# Development and clinical application of a methylated ctDNA assay in the preoperative risk classification of resectable colon cancer

**DOI:** 10.1101/2024.12.04.24318421

**Authors:** L Raunkilde, RF Andersen, S Timm, M Jørgensen, J Lindebjerg, SR Rafaelsen, TF Hansen, LH Jensen

## Abstract

**Background:** Pathological lymph node involvement (pN+) is an indicator of poor prognosis in localized colon cancer. Currently, the preoperative risk classification fails to accurately identify pN+, which could be instrumental in the selection of patients for neoadjuvant chemotherapy (NAC). This study aimed to develop an assay for preoperative methylated circulating tumor DNA (meth-ctDNA) as a predictive marker of pN+ and as a biomarker for initiating NAC in patients with localized colon cancer.

**Materials and methods:** 203 patients operated for colon cancer stage I-III at Vejle Hospital were randomly assigned (1:1) to a discovery and a validation cohort. A multiplex assay of a tumor-specific (NPY) and two organ-specific (GAL3ST3, KANK1) meth-ctDNAs was designed and methylation analysis was performed using droplet digital PCR. The ability of preoperative meth-ctDNA to predict pN+ was assessed using receiver operating characteristics (ROC) analysis.

**Results:** ROC analyses of meth-ctDNA (negative/positive) to predict pN+ had an area under the curve of 52% and 57% in the discovery and validation cohort, respectively. Survival analysis in the validation cohort confirmed a higher rate of 48 months disease-free survival in meth-ctDNA negative compared to positive patients (HR=2.31, 95% CI 1.01-5.2). The difference also applied to 4-year overall survival (HR=3.62, 95% CI 1.15-11.4). In a multivariate analysis, meth-ctDNA remained the strongest prognostic factor of DFS (HR 2.46, 95% CI 1.03-5.88) and OS (HR 4.54, 95% CI 1.32-15.6) in relation to clinical T- or N-category, gender and age.

**Conclusion:** The preoperative meth-ctDNA multiplex assay did not predict pN+. The findings suggest meth-ctDNA as a more powerful marker than N-category in identifying high-risk patients who would potentially benefit from NAC.

## Introduction

In 2020, GLOBOCAN estimated the worldwide number of new colon cancer cases to be 1,148,515^1^. Morbidity and mortality are significant, and the overall 5-year survival rate is around 60%^2^. Approximately 80% of colon cancer patients present with localized disease and can undergo surgery with curative intent. Postoperative adjuvant chemotherapy (AC) is standard in localized colon cancer patients with pathological lymph node involvement (pN+) and in stage II patients with specific risk factors, many of whom are over-treated^3,4^.

Neoadjuvant chemotherapy (NAC) prior to surgery may have potential advantages over postoperative AC. Tumor shrinkage may reduce the risk of incomplete resection and tumor-shedding during surgery^5^, and systemic therapy could potentially reduce the presence of micrometastasis weeks before postoperative AC^6^ and thereby improve survival. Currently, NAC is not the standard of treatment in operable colon cancer patients partly due to concerns about the accuracy of radiological tumor staging^7^. There is a clear need for biomarkers that allow a more personalized preoperative risk classification for localized disease than is currently possible with the clinical (c) tumor, node, and metastasis (TNM) classification of colon cancer. This classification, as outlined in Brierley 2017, 8^th^ edition, is based on an overall assessment of clinical, endoscopic, and radiological findings before treatment.

In all guidelines, pN+ is recognized as an indicator of poor prognosis in colon cancer, typically prompting the use of postoperative AC. If pN+ could be reliably predicted prior to surgery, it could serve as a biomarker for the initiation of NAC.

Circulating tumor-specific DNA (ctDNA) obtained by a liquid biopsy represents a relatively new biomarker that can be measured as a fraction of circulating total cell-free DNA (cfDNA). Droplet digital polymerase chain reaction (ddPCR) has high sensitivity and allows for detection of very small fractions of ctDNA^8,9^. Aberrant methylation patterns occur in most malignant tumors and distinct, cancer-specific DNA hypermethylation in the regulatory regions of specific genes is a potential biomarker measurable in plasma as methylated tumor specific DNA (meth-ctDNA). Hypermethylation of the *neuropeptide Y gene* (*NPY* meth-ctDNA) occurs in most colorectal cancers but not in normal colorectal tissue^10^. Furthermore, organ-specific markers are investigated as potential markers of cancer^11^.

The objective of this study was to develop a meth-ctDNA assay and subsequently assess the utility of preoperative meth-ctDNA as a predictive marker for pN+ and to evaluate its potential clinical value as a biomarker for initiating NAC.

## Material and methods

The study was approved by the Regional Committee on Health Research Ethics for Southern Denmark (S-20190181). Written and orally informed consent to translational research according to the Helsinki II Declaration was obtained from all patients.

The reporting of the study follows the REMARK guideline^12^

### Patient eligibility

The clinical data were collected from the Colorectal Cancer Database established at Danish Colorectal Cancer Center South, Vejle Hospital, Denmark.

Data were extracted consecutively according to the inclusion criteria; 1) consent to inclusion in the Colorectal Cancer Database and to blood sampling; 2) the ICD 10 classification code for colon cancer 3) biopsy proven adenocarcinoma 4) upfront surgery 5) blood sampled before surgery; 6) clinical TNM stage I-III disease (T1-T4, N0-N2, M0). Patients were manually excluded if they were treated with NAC or developed metastatic disease (synchronous metastases) within six months from surgery. Malignant disease within three years before surgery for colon cancer, except basal cell or squamous cell carcinoma of the skin or carcinoma in situ cervicis uteri, was an exclusion criterion.

Clinical data from the Colorectal Cancer Database were transferred to and managed using the Research Electronic Data Capture (REDCap) tool^13,14^ hosted by the Odense Patient Data Explorative Network (OPEN). Patients were included and randomized into two cohorts (discovery and validation) using the electronic randomization module OPEN Randomize.

### Identification of colon specific methylation sites using Illumina 450K methylation arrays

The data used for this analysis consisted of 450K methylation data for all 341 colon adenocarcinoma (COAD) samples in The Cancer Genome Atlas (TCGA) along with all normal samples (n=708) for all available solid tumor types (n=22) in the TCGA as of April 2020. For details, please see Supplemental Table 1. In addition, selected previously published and publicly available datasets were used: GSE67393 (peripheral blood, n=117)^15^, GSE121192 (peripheral blood monocytes, n=16)^16^, and GSE50874 (kidney tissue from healthy subjects, n=85)^17^. Finally, an in-house dataset consisting of 57 colon samples was used (healthy normal/ non-cancerous colon tissue, n=34), and patients with colon polyps or colon cancer, n=23). In total, the dataset included 1,324 samples (926 non-colon normal samples and 398 samples from colon).

All data analyses were performed in R (v. 4.0.5, https://www.r-project.org/ ) using the packages caret (v.6.0-92), randomForest (v. 4.6-14), mlbench (v. 2.1-3), pheatmap (v. 1.0.12), ggplot2 (v.3.3.5), dplyr (v. 1.0.7), readr (v.2.1.2) and minfi in addition to functions in base packages.

Initially, methylation probes with missing values were filtered out. Mean beta-values for all colon samples (COAD) and all non-colon samples (REST) were calculated. An initial list of differentially methylated CpGs (DMC) was extracted by selecting sites with mean COAD beta > 0.75 and mean REST beta <0.25 (i.e. colon hypermethylated sites) or mean COAD beta <0.25 and mean REST beta >0.75 (i.e. colon hypomethylated sites). A total of 123 DMCs for 1,324 samples passed this initial analysis. We omitted samples from the TCGA Rectum Adenocarcinoma group, as it shows highly similar methylation patterns as the COAD samples. We then used recursive feature elimination (RFE) using 10-fold cross-validation (repeated five times) to identify the DMCs best suited to separate colon from non-colon samples. Similarly, we used RandomForest (10 fold cross-validation, repeated 10 times) to detect the most important variables (DMCs) that contributed the most to separating colon from non-colon samples. Finally, we merged the resulting top 10 DMC lists from both the RFE and the random forest analysis. In total, 11 unique DMCs were identified. Following manual inspection of the beta value distribution for each sample group, i.e. each TCGA tumor type or GSE dataset, we discarded three DMCs that were considered inappropriate for further analysis. In total, a list of eight unique DMCs were used as candidates for further investigation.

### Assay development

Methylation of the *NPY* ^18^ was chosen as a colon cancer-specific marker based on previous work^10^. ddPCR assays were designed for regions around the eight unique DMCs using the PrimerQuest Tool (https://eu.idtdna.com/Primerquest/Home/Index) and initial testing was done on tissue samples (colon and rectum tumor and non-cancerous tissue), genomic DNA from whole blood, and plasma from healthy individuals. Assays were discarded for various reasons: not possible to design efficient ddPCR assay, unspecific signal when analyzing genomic DNA from whole blood, marker was positive in plasma from healthy individuals, and low signal in non-cancerous tissue. Two assays passed initial testing (KANK1, KN Motif And Ankyrin Repeat Domains 1; GAL3ST3, Galactose-3-O-Sulfotransferase 3) and were included in a multiplex as described below.

### Colon multiplex assay

Sample and mastermix were combined in 48 µl reaction volume according to Supplemental Table 2. 22 µl were distributed into two wells for droplet generation. Supplemental Figure 1 shows the fluorescence plots representing a positive and a negative plasma sample.

### Assay validation

The multiplex assay was tested on DNA from colon and rectal non-cancerous tissue and colon and rectum tumor tissue. The samples were formalin fixed and paraffin embedded. DNA was extracted using the Maxwell extraction system (Promega, Madison, WI, USA).

DNA from tumor tissue was analyzed with single assays for the three individual markers and with the multiplex assay. The non-tumor samples were only tested with multiplex analysis. Supplemental Figure 2 shows the percentage of the three markers measured by singleplex and multiplex analysis in colon and rectal tumor tissue.

### Analysis of meth-ctDNA

A blood sample was collected in EDTA tubes before surgery and plasma was isolated by centrifugation (2,000g for 10 min) within four hours of sampling. Plasma was stored at −80 °C until analysis. After thawing, plasma was centrifuged again at 10,000 g for 10 min and Cysteine-rich polycomb-like protein1 (CPP1) was added as exogenous internal control ^19^. DNA was extracted from 4 mL of plasma with the QIAsymphony purification system and QIAsymphony Circulating DNA kit (Qiagen, Hilden, Germany) according to the manufacturer’s instructions. 340 ul water was added and samples were analyzed by qPCR to quantify CPP1 and total cell free DNA (gB2M) ^19^ using 3 ul of DNA per well. DNA was concentrated to 20 μl on Amicon Ultra Centrifugal filter units (Millipore). The concentrated DNA was bisulfite converted in a 150 μl reaction with EZ DNA Methylation Lightning kit (Zymo Research, Irvine, CA, USA). DNA was eluted in 15 μl Universal human methylated control DNA (Zymo Research, Irvine, CA, USA), genomic DNA from whole blood and water were converted alongside the samples as positive and negative controls.

For methylation analysis, droplet digital PCR was used. Multiplex analysis was run in duplicate with 5 μl of DNA per well in a 20 μl reaction. Droplet digital polymerase chain reaction multiplex supermix for probes (no dUTP) was used.

Droplets were made in the automated droplet generator from BioRad (BioRad, Hercules, CA, USA). ddPCR was run on PCR equipment Veriti Thermal Cycler. The samples were read on the QX200 Droplet Reader from BioRad. Data analysis was performed with QX Manager Software 2.0. (BioRad).

The methylation analysis of the respective genes, NPY, GAL3ST3 and KANK1, was reported as number of droplets per analysis, number of DNA copies (meth-ctDNA copies/ml plasma), and as a percentage of total DNA ((meth-ctDNA copies/albumin copies) × 100). The methylation analysis was performed blinded to the study endpoints.

### Endpoints

The primary endpoint was the ability of preoperative meth-ctDNA to predict the risk of pN+. Secondary endpoints were the association between meth-ctDNA and disease free survival (DFS) and overall survival (OS), respectively, and between preoperative meth-ctDNA and pathological risk factors preoperatively as well as postoperatively.

### Statistical analysis

Analysis of the ability of preoperative meth-ctDNA to predict pN+ was based on receiver operating characteristics (ROC). Specifically, the ability of the mean, sum and maximum of the percentage and copy numbers of three ctDNAs, respectively, to predict pN+ was presented as ROC curves. An area under the ROC curve (AUC) of ≥ 70% was deemed of interest. Meth-ctDNA as a dichotomized variable (negative/positive) was investigated as a diagnostic test to predict pN+ based on ROC analysis. The diagnostic performance of the preoperative CT scan with respect to pN+ was also elucidated by ROC analysis. The sample size was based on meth-ctDNA being superior to cN+ evaluated by CT scan in predicting pN+. To detect a 20% difference from the random value (50%) approximately 85 patients were to be enrolled in each cohort with a power of 90% and a risk of type 1 error of 5%. To allow for up to 15% missing data, at least 200 patients were to be be enrolled and randomized to the discovery or validation cohort.

The association between meth-ctDNA and a number of suggested risk factors (pre- and postoperative T- and N-category, V-(vascular)category and perineural invasion, tumor budding, mismatch repair status) was investigated by Spearman’s rank coefficient analysis, Wilcoxon rank sum test, Pearson Chi-squared test or Fishers Exact test where applicable. DFS and OS according to the status of meth-ctDNA were based on the Kaplan-Meier estimator and compared by the log rank test. Independent prognostic impact was tested in multiple Cox regression analysis. The parameters entered in the multivariate Cox model were conventional covariates such as age and gender in addition to preoperative variables with a p-value lower than 0.1 with any of the time-to-event endpoints in the univariate Cox analysis (T-category, N-category). The assumption on proportional hazards was assessed by log-log plots, observed vs. estimated survival curves, and the Schoenfeld residuals, and they were accepted.

Statistical calculations were performed using Stata/BE 18.0 (StataCorp LLC, TX, USA).

## Results

We enrolled 260 patients from the Colorectal Cancer Database who had undergone surgical treatment for colon cancer between January 1, 2017 and August 1, 2019 and met the inclusion criteria. Among the enrolled patients, 57 were excluded based on the exclusion criteria, resulting in a final sample size of 203 patients. Of these, 102 and 101 patients were randomized to the discovery and validation cohort, respectively. For details, please see flow chart in Supplemental Figure 3. The data cut-off for survival analysis was set at 1 June 2022 to allow for at least three years of follow-up.

Based on analysis of the discovery cohort samples were classified as positive if two out of three methylation markers had ≥ three positive droplets.

The baseline characteristics in the two cohorts were comparable (Table 1). In the discovery cohort, 41 patients (40%) were positive for meth-ctDNA compared to 46 patients (46%) in the validation cohort. The prevalence of pN+ in the discovery and validation cohorts was 32% and 35%, respectively, with 44% and 54% of the cases testing positive for meth-ctDNA. In the discovery and validation cohorts 56% and 54%, respectively, received postoperative AC. None of the patients with stage II cancer in the discovery cohort received postoperative AC compared to four patients (9%) in the validation cohort.

**Table 1.**
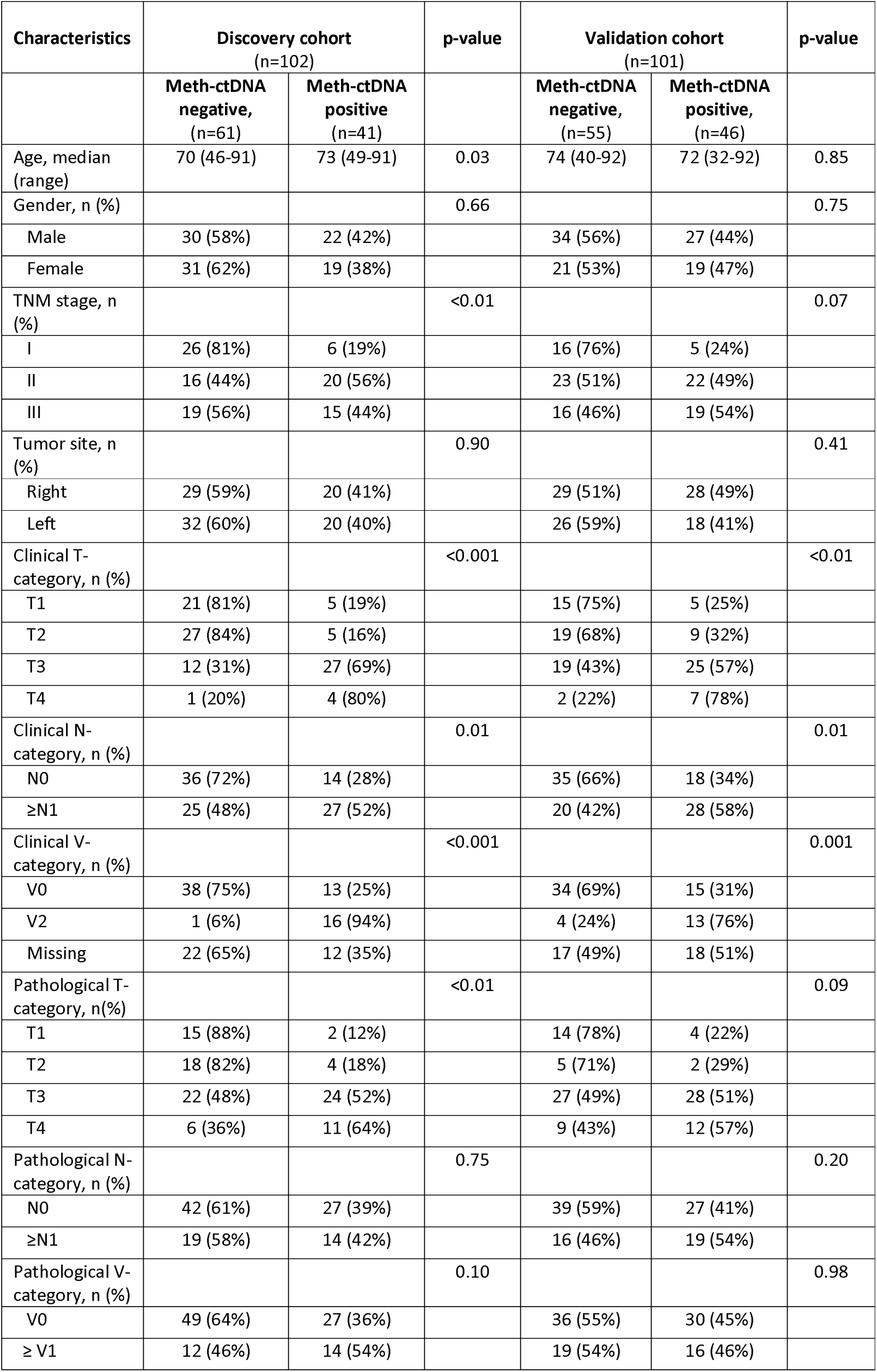
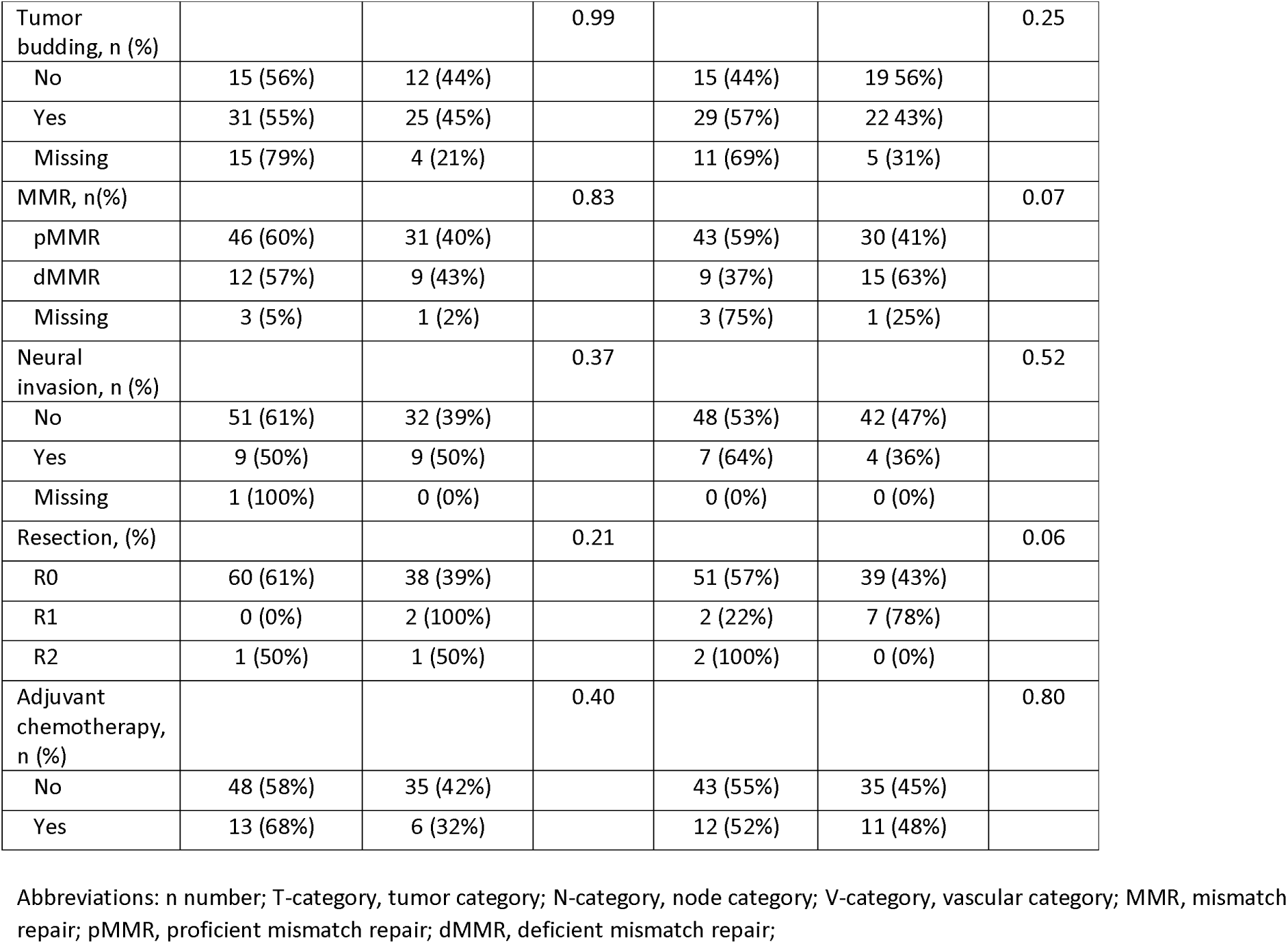
Patient characteristics at baseline and relation to meth-ctDNA.

| Characteristics | Discovery cohort<br>(n=102) |  | p-value | Validation cohort<br>(n=101) |  | p-value |
| --- | --- | --- | --- | --- | --- | --- |
|  | Meth-ctDNA<br>negative,<br>(n=61) | Meth-ctDNA<br>positive<br>(n=41) |  | Meth-ctDNA<br>negative,<br>(n=55) | Meth-ctDNA<br>positive,<br>(n=46) |  |
| Age, median<br>(range) | 70 (46-91) | 73 (49-91) | 0.03 | 74 (40-92) | 72 (32-92) | 0.85 |
| Gender, n (%) |  |  | 0.66 |  |  | 0.75 |
| Male | 30 (58%) | 22 (42%) |  | 34 (56%) | 27 (44%) |  |
| Female | 31 (62%) | 19 (38%) |  | 21 (53%) | 19 (47%) |  |
| TNM stage, n<br>(%) |  |  | <0.01 |  |  | 0.07 |
| I | 26 (81%) | 6 (19%) |  | 16 (76%) | 5 (24%) |  |
| II | 16 (44%) | 20 (56%) |  | 23 (51%) | 22 (49%) |  |
| III | 19 (56%) | 15 (44%) |  | 16 (46%) | 19 (54%) |  |
| Tumor site, n<br>(%) |  |  | 0.90 |  |  | 0.41 |
| Right | 29 (59%) | 20 (41%) |  | 29 (51%) | 28 (49%) |  |
| Left | 32 (60%) | 20 (40%) |  | 26 (59%) | 18 (41%) |  |
| Clinical T-category, n (%) |  |  | <0.001 |  |  | <0.01 |
| T1 | 21 (81%) | 5 (19%) |  | 15 (75%) | 5 (25%) |  |
| T2 | 27 (84%) | 5 (16%) |  | 19 (68%) | 9 (32%) |  |
| T3 | 12 (31%) | 27 (69%) |  | 19 (43%) | 25 (57%) |  |
| T4 | 1 (20%) | 4 (80%) |  | 2 (22%) | 7 (78%) |  |
| Clinical N-category, n (%) |  |  | 0.01 |  |  | 0.01 |
| N0 | 36 (72%) | 14 (28%) |  | 35 (66%) | 18 (34%) |  |
| ≥N1 | 25 (48%) | 27 (52%) |  | 20 (42%) | 28 (58%) |  |
| Clinical V-category, n (%) |  |  | <0.001 |  |  | 0.001 |
| V0 | 38 (75%) | 13 (25%) |  | 34 (69%) | 15 (31%) |  |
| V2 | 1 (6%) | 16 (94%) |  | 4 (24%) | 13 (76%) |  |
| Missing | 22 (65%) | 12 (35%) |  | 17 (49%) | 18 (51%) |  |
| Pathological T-category, n(%) |  |  | <0.01 |  |  | 0.09 |
| T1 | 15 (88%) | 2 (12%) |  | 14 (78%) | 4 (22%) |  |
| T2 | 18 (82%) | 4 (18%) |  | 5 (71%) | 2 (29%) |  |
| T3 | 22 (48%) | 24 (52%) |  | 27 (49%) | 28 (51%) |  |
| T4 | 6 (36%) | 11 (64%) |  | 9 (43%) | 12 (57%) |  |
| Pathological N-category, n (%) |  |  | 0.75 |  |  | 0.20 |
| N0 | 42 (61%) | 27 (39%) |  | 39 (59%) | 27 (41%) |  |
| ≥N1 | 19 (58%) | 14 (42%) |  | 16 (46%) | 19 (54%) |  |
| Pathological V-category, n (%) |  |  | 0.10 |  |  | 0.98 |
| V0 | 49 (64%) | 27 (36%) |  | 36 (55%) | 30 (45%) |  |
| ≥ V1 | 12 (46%) | 14 (54%) |  | 19 (54%) | 16 (46%) |  |
| Tumor budding, n (%) |  |  | 0.99 |  |  | 0.25 |
| No | 15 (56%) | 12 (44%) |  | 15 (44%) | 19 (56%) |  |
| Yes | 31 (55%) | 25 (45%) |  | 29 (57%) | 22 (43%) |  |
| Missing | 15 (79%) | 4 (21%) |  | 11 (69%) | 5 (31%) |  |
| MMR, n(%) |  |  | 0.83 |  |  | 0.07 |
| pMMR | 46 (60%) | 31 (40%) |  | 43 (59%) | 30 (41%) |  |
| dMMR | 12 (57%) | 9 (43%) |  | 9 (37%) | 15 (63%) |  |
| Missing | 3 (5%) | 1 (2%) |  | 3 (75%) | 1 (25%) |  |
| Neural invasion, n (%) |  |  | 0.37 |  |  | 0.52 |
| No | 51 (61%) | 32 (39%) |  | 48 (53%) | 42 (47%) |  |
| Yes | 9 (50%) | 9 (50%) |  | 7 (64%) | 4 (36%) |  |
| Missing | 1 (100%) | 0 (0%) |  | 0 (0%) | 0 (0%) |  |
| Resection, (%) |  |  | 0.21 |  |  | 0.06 |
| R0 | 60 (61%) | 38 (39%) |  | 51 (57%) | 39 (43%) |  |
| R1 | 0 (0%) | 2 (100%) |  | 2 (22%) | 7 (78%) |  |
| R2 | 1 (50%) | 1 (50%) |  | 2 (100%) | 0 (0%) |  |
| Adjuvant chemotherapy, n (%) |  |  | 0.40 |  |  | 0.80 |
| No | 48 (58%) | 35 (42%) |  | 43 (55%) | 35 (45%) |  |
| Yes | 13 (68%) | 6 (32%) |  | 12 (52%) | 11 (48%) |  |

Abbreviations: n number; T-category, tumor category; N-category, node category; V-category, vascular category; MMR, mismatch repair; pMMR, proficient mismatch repair; dMMR, deficient mismatch repair;

### Prediction of postoperative lymph node metastases

Investigating the ability of the mean, sum and maximum of the percentage and copy numbers of the three meth-ctDNAs to predict pN+, the AUCs were around 50% in the discovery cohort. In the validation cohort the AUCs were around 60%. Hence, none of the cohorts approached the 70% that was of interest (Supplemental Figure 4). The sensitivity and specificity of meth-ctDNA was low (42% and 61%, respectively) in the discovery cohort when analyzed as a diagnostic test to predict pN+ (positive/negative). The AUC was 52% and the positive and negative predictive value was 34% and 69%, respectively. Similar results were seen in the validation cohort (Table 2). The diagnostic performance of the preoperative CT scan (cN) in predicting pN+ showed AUC values of 57% and 60% in the discovery and validation cohort, respectively.

**Table 2.** ROC analysis of dichotomized meth-ctDNA as a diagnostic test to predict pN+.

|  | **Discovery cohort (95% CI)** | **Validation cohort (95% CI)** |
| --- | --- | --- |
| Prevalence pN+ | 32% | 35% |
| Sensitivity | 42% (26-61) | 54% (37-71) |
| Specificity | 61% (48-72) | 59% (46-71) |
| AUC | 52% (41-62) | 57% (46-67) |
| PPV | 34% (20-51) | 41% (27-57) |
| NPV | 69% (56-80) | 71% (57-82) |

Abbreviations: pN+, pathological lymph node; AUC, area under the curve; PPV, positive predictive value; NPV, negative predictive value

### Associations and correlations

In the discovery cohort preoperative meth-ctDNA was correlated to cT (rho=0.48, p<0.001) and pT (rho=0.40, p<0.01). Also, in the validation cohort meth-ctDNA was correlated to cT (rho=0.33, p<0.01) and pT (rho=0.23, p=0.09). In both cohorts meth-ctDNA was associated to cN (p=0.01). A strong association was found between meth-ctDNA and the clinical V2-category in both cohorts (p<0.001 and p=0.001, respectively), although more than 30% of the patients in each cohort had a missing value in this specific variable. There was no association between meth-ctDNA and pathological vascular invasion in any of the cohorts. The same applies between meth-ctDNA and perineural invasion, pathological venous invasion, tumor budding, mismatch repair, grade of resection, and postoperative AC. In the discovery cohort preoperative meth-ctDNA was associated with TNM stage (p<0.01), but in the validation cohort the association was not statistically significant (p=0.07) (Table 1).

### Preoperative meth-ctDNA and survival

The median follow-up time was comparable in the two cohorts; 49.1 months (95% CI 44.9-51.7) and 49.5 months (95% CI 46.0-52.7), respectively. Figure 1a-d illustrates the DFS and OS in patients according to their preoperative meth-ctDNA status. After 48 months, the DFS was 89.7% in patients with negative meth-ctDNA in the discovery cohort compared to 59.8% in patients with positive meth-ctDNA (HR=3.07, 95% CI 1.21-7.82, p=0.01). Similarly, in the validation cohort, the 48-month DFS was 77.3% in patients with negative meth-ctDNA compared to 62.4% in patients with positive meth-ctDNA (HR=2.31, 95% CI 1.02-5.22, p=0.04). These differences were also observed in the 4-year OS. In the discovery cohort 95.0% of meth-ctDNA negative patients were still alive compared to 75.0% of the patients with positive meth-ctDNA (HR=5.19, 95% CI 1.40-19.20, p<0.01). In the validation cohort 91.0% of meth-ctDNA negative patients were alive after four years compared to 78.0% of the meth-ctDNA positive patients (HR=3.62, 95% CI 1.15-11.38, p=0.02).

**Figure 1.**
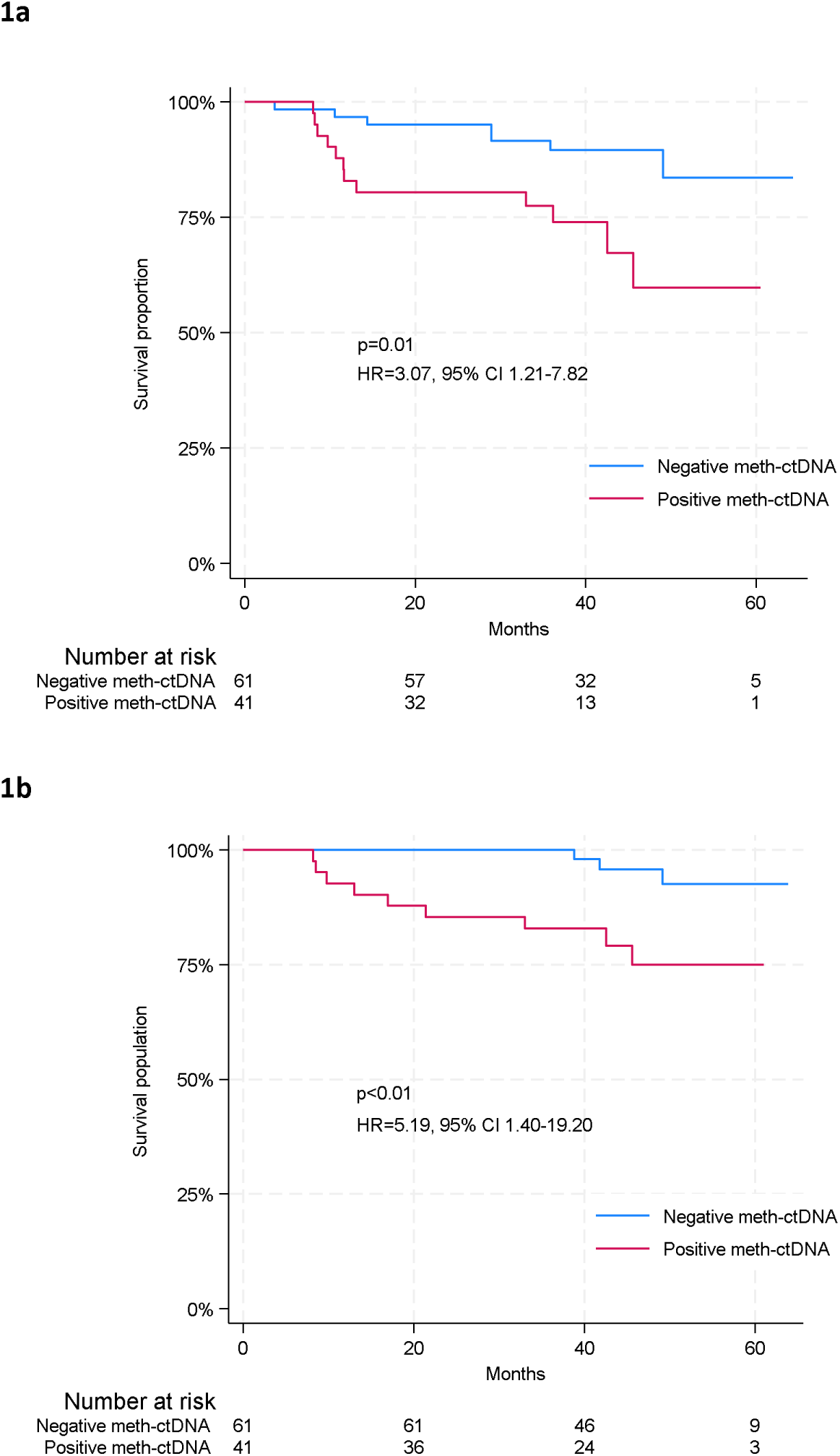
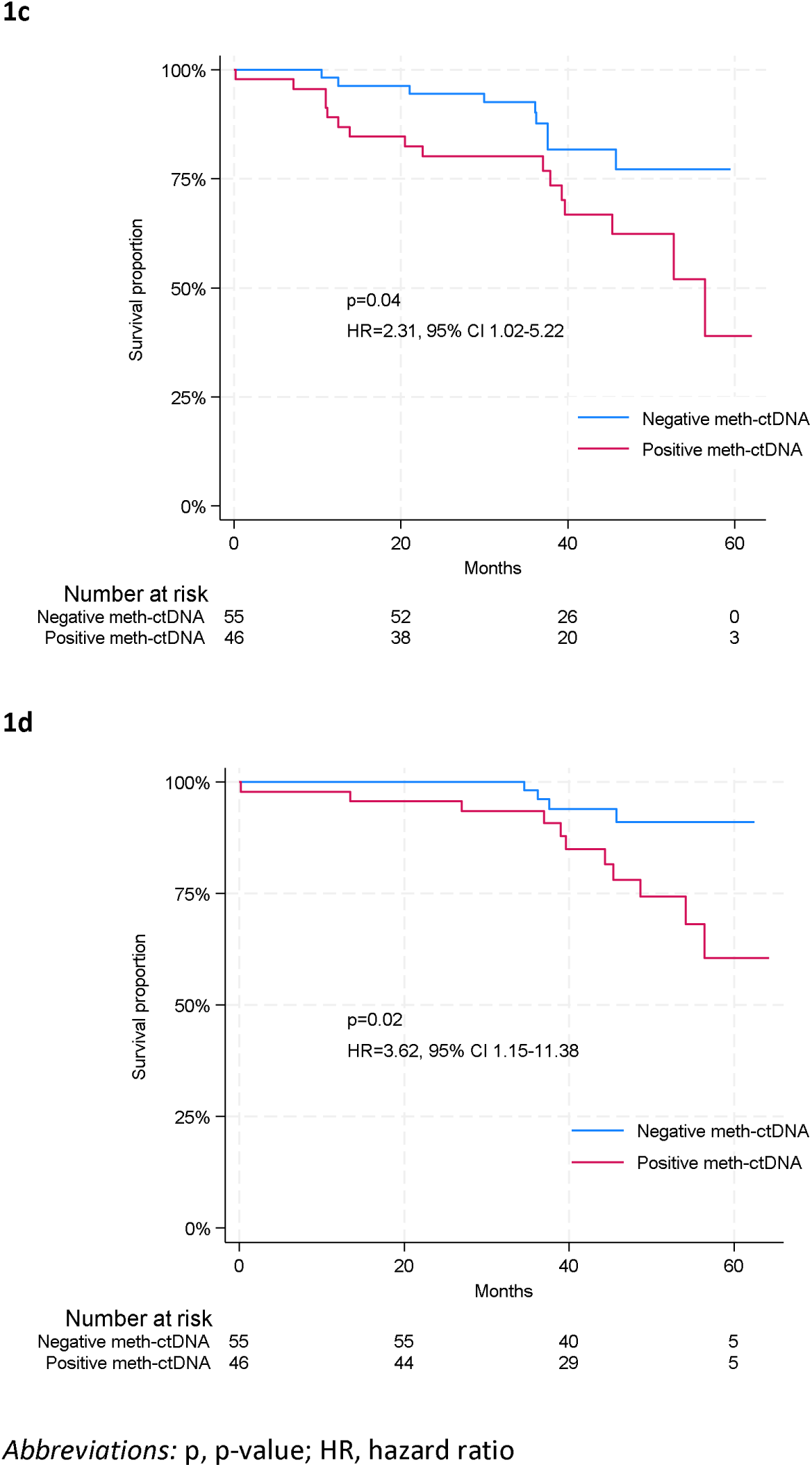
Kaplan-Meier plots describing (a) disease-free survival in the discovery cohort according to meth-ctDNA, (b) overall survival in the discovery cohort according to meth-ctDNA, (c) disease-free survival in the validation cohort according to meth-ctDNA, (d) overall survival in the validation cohort according to methctDNA

In a multivariate analysis of the validation cohort, meth-ctDNA remained the strongest prognostic factor of DFS (HR 2.25, 95% CI 0.95-5.30, p=0.07) and OS (HR 4.35, 95% CI 1.31--14.42, p=0.02) compared to clinical T- or N-category, gender and age (Table 3).

**Table 3.** Multiple Cox regression analysis of survival in the validation cohort.

| **Variable** | **DFS**<br>**HR (95% CI)** | **p-value** | **OS**<br>**HR (95% CI)** | **p-value** |
| --- | --- | --- | --- | --- |
| Meth-ctDNA<br>Undetectable<br>Detectable | Reference<br>2.25 (0.95-5.30) | 0.07 | Reference<br>4.35 (1.31-14.42) | 0.02 |
| Age | 0.98 (0.94-1.02) | 0.28 | 1.02 (0.97-1.07) | 0.51 |
| Gender<br>Male<br>Female | Reference<br>0.60 (0.26-1.40) | 0.24 | Reference<br>0.67 (0.23-1.98) | 0.47 |
| cT category<br>cT1-T2<br>cT3-T4 | Reference<br>1.06 (0.43-2.63) | 0.90 | Reference<br>0.47 (0.15-1.43) | 0.18 |
| cN category<br>cN0<br>cN≥1 | Reference<br>1.17 (0.50-2.77) | 0.71 | Reference<br>1.42 (0.47-4.28) | 0.54 |

Abbreviations: DFS, disease-free survival; OS, overall survival; HR, hazard ratio; CI, confidence interval; cT, clinical tumor; cN, clinical node

**OS**
| **Variable** | **HR (95% CI)** | **p-value** |
| --- | --- | --- |
| Meth-ctDNA<br>Undetectable<br>Detectable | Reference<br>4.35 (1.31-14.42) | 0.02 |
| Age | 1.02 (0.97-1.07) | 0.51 |
| Gender<br>Male<br>Female | Reference<br>0.67 (0.23-1.98) | 0.47 |
| cT category<br>cT1-T2<br>cT3-T4 | Reference<br>0.47 (0.15-1.43) | 0.18 |
| cN category<br>cN0<br>cN≥1 | Reference<br>1.42 (0.47-4.28) | 0.54 |

Abbreviations: DFS, disease-free survival; OS, overall survival; HR, hazard ratio; CI, confidence interval; cT, clinical tumor; cN, clinical node

A post hoc analysis of pooled cohorts demonstrated a statistically significant difference in DFS with an HR of 2.78 (95% CI 1.50-5.15, p<0.001) in favor of the meth-ctDNA negative patients. The difference also applied to OS with an HR of 4.42 (95% CI 1.87-10.48, p<0.001). In a pooled multivariate analysis meth-ctDNA remained the strongest prognostic factor compared to clinical T- or N-category, gender and age for both DFS (HR 2.08, 95% CI 1.06-4.06, p=0.03) and OS (HR 3.89, 95% CI 1.56-9.75, p<0.01) (Table 4).

**Table 4.** Multiple Cox regression analysis of survival in the pooled cohort.

| **Variable** | **DFS**<br>**HR (95% CI)** | **p-value** | **OS**<br>**HR (95% CI)** | **p-value** |
| --- | --- | --- | --- | --- |
| Meth-ctDNA<br>Undetectable<br>Detectable | Reference<br>2.08 (1.06-4.06) | 0.03 | Reference<br>3.89 (1.56-9.75) | <0.01 |
| Age | 1.00 (0.97-1.04) | 0.77 | 1.04 (1.00-1.09) | 0.04 |
| Gender<br>Male<br>Female | Reference<br>0.61 (0.33-1.15) | 0.13 | Reference<br>0.49 (0.21-1.12) | 0.09 |
| cT category<br>cT1-T2<br>cT3-T4 | Reference<br>1.70 (0.80-3.59) | 0.17 | Reference<br>1.27 (0.50-3.24) | 0.62 |
| cN category<br>cN0<br>cN≥1 | Reference<br>1.30 (0.66-2.55) | 0.44 | Reference<br>1.13 (0.50-2.59) | 0.76 |

**OS**
| **Variable** | **HR (95% CI)** | **p-value** |
| --- | --- | --- |
| Meth-ctDNA<br>Undetectable<br>Detectable | Reference<br>3.89 (1.56-9.75) | <0.01 |
| Age | 1.04 (1.00-1.09) | 0.04 |
| Gender<br>Male<br>Female | Reference<br>0.49 (0.21-1.12) | 0.09 |
| cT category<br>cT1-T2<br>cT3-T4 | Reference<br>1.27 (0.50-3.24) | 0.62 |
| cN category<br>cN0<br>cN≥1 | Reference<br>1.13 (0.50-2.59) | 0.76 |

Abbreviations: cT, clinical tumor; cN, clinical node

## Discussion

In this retrospective-prospective biomarker study on patients with resectable colon cancer, preoperative meth-ctDNA did not predict pN+. It was, however, a strong prognostic marker as confirmed in the validation cohort. Meth-ctDNA positive patients had a significantly shorter DFS and OS compared to meth-ctDNA negative patients. A pooled multivariate analysis further confirmed preoperative meth-ctDNA as the strongest prognostic factor for both DFS (HR 2.08, 95% CI 1.06-4.06) and OS (HR 3.89, 95% CI 1.56-9.75). Specifically, meth-ctDNA proved better than N-category in predicting DFS and OS. Interestingly, meth-ctDNA was detected in 50% of the patients with stage II disease, a group of patients for whom better selection criteria for postoperative AC are needed. It should be noted that our study was not powered to do subgroup analyses on meth-ctDNA in different stages and survival.

We have presented a new method by combining a tumor-specific marker and two organ-specific markers in a methylation analysis. Organ-specific markers are of interest based on the principles that dying cells release cfDNA and each type of tissue has a specific DNA methylation pattern^20^. Moss et al. have proposed breast-specific methylated cfDNA as a universal marker for detection of breast cancer^21^, and Gai et al. found increased levels of colon-derived DNA in patients with CRC compared to healthy controls^22^. The three specific markers in the present multiplex were chosen on a technical basis and a well-performing assay with the expectation of increased sensitivity.

In a methylation analysis, every patient is analyzed with the same method. One strength in the methylation analysis is the universal character of the marker, which is typically more frequently observed than mutations. Also, the hypermethylation of a gene is usually constant in the tumor cells. A disadvantage of the method is increasing cfDNA by age^23^. In addition, there is a risk of damage or loss of DNA during bisulfite conversion, which reduces the overall sensitivity of the assay.

In the present study, several patients in stage I and II were meth-ctDNA positive. This was also one of the findings by Jensen et al. who investigated three tumor-specific DNA methylation markers for detection of localized CRC^24^.

Recently, Bregni et al. have published the first retrospective study of meth-ctDNA (NPY and WIF1) in the neoadjuvant setting of stage III colon cancer receiving one cycle of FOLFOX^25^. They found that 25 of 60 patients (42%) had preoperative, detectable meth-ctDNA, which is comparable to our results (44% and 54% of stage III in the discovery and validation cohort, respectively). However, their definition of ‘detectable’ was presence of either ≥ 2 droplets of NPY or ≥ 5 droplets of WIF1, which is different from our definition. They also found that circulating free DNA, but not meth-ctDNA as in our study, was an independent prognostic factor. The detection rates of meth-ctDNA in our study also support the rates by Garrigou et al.^26^, and as expected the rate is lower than in metastatic tumors^27,28^. Bregni and Garrigou are both investigating NPY and WIF1. There are differences in methods and hence cut-off compared to our present report. Such differences contribute to the diversity in reporting and complicate implementation^29^.

During recent years, the research in ctDNA has focused on minimal residual disease in the postoperative setting of early stage or metastatic tumors ^30,31,32^ and treatment monitoring in the palliative setting of CRC ^33,34^. Few studies have centered on the neoadjuvant setting, where the challenge is to identify the specific marker (or markers) that predicts effect of NAC or a poor prognosis. NAC has the potential to improve the outcome of selected groups of colon cancer patients, but selection criteria have not been defined. Historically, lymph node involvement has been an important risk factor of recurrence, but 50% of patients with stage III colon cancer are cured with surgery alone^35^. ctDNA in plasma suggests more advanced disease, but whether it can stand alone as a risk marker remains to be proven. A future re-assessment of the preoperative risk classification in colon cancer must be considered.

The FOxTROT trial was the first randomized controlled trial investigating NAC with three cycles of 5-fluorouracil and oxaliplatin in locally advanced colon cancer patients^36^. The inclusion criteria were T-stage 3-4 with extramural extension ≥ 1 mm, N-stage 0-2, M-stage 0, equivalent to stage II and III. It showed fewer recurrences at 2 years among patients who received NAC compared to patients in the control arm receiving postoperative AC (p=0.042). A number of these patients are overtreated, and a biomarker for better selection of patients is the key for personalizing NAC in colon cancer. Results are eagerly awaited on minimal residual disease by ctDNA and ctDNA alterations during NAC from the currently recruiting FOxTROT-2 and FOxTROT-3 studies investigating dose-modified NAC in elderly and dose-escalation in younger people, respectively^37^.

In the FOxTROT trial, patients with deficient mismatch repair (dMMR) tumors did not benefit from NAC. In our study, there was no difference in meth-ctDNA between patients with dMMR and patients with proficient mismatch repair (pMMR) tumors. Data from the NICHE-2 trial on stage II-III colon cancer patients with dMMR tumors receiving neoadjuvant check point inhibitors showed major pathological response in 95% of the patients^38^, suggesting this treatment has potential to become standard of care in the near future. For this subgroup of colon cancer patients dMMR may be the marker for NAC.

The strengths in our study are the use of discovery and validation cohorts with patients and samples prospectively collected at the time of diagnosis. Additionally, the study included new organ-specific markers and the analyses were all performed in the same laboratory. Limitations include a sample size not powered for subgroup analysis and laboratory analyses performed retrospectively on stored samples, which could have affected the amount of DNA. Also, the impact of NAC or postoperative AC was not evaluable in this cohort. Taking this into account, our study represents a step towards a re-evaluation of the preoperative risk classification of colon cancer and the clinical utility of ctDNA biomarkers in the neoadjuvant setting.

## Conclusions

We have developed and clinically tested an assay for meth-ctDNA in colon cancer. Although preoperative detection of meth-ctDNA was not associated with pathological lymph node involvement, the results showed that it serves as a potent prognostic marker. This suggests that preoperative meth-ctDNA may be more powerful than N-category in identifying high-risk patients who will potentially benefit from NAC. We recommend initiation of randomized controlled trials to assess the efficacy of NAC in colon cancer patients with positive meth-ctDNA preoperatively.

### Clinical Practice Points

- The current preoperative risk classification in colon cancer falls short in accurately identifying patients with pathological lymph node involvement. A more personalized approach is required to identify patients who would benefit from neoadjuvant treatment (NAC).
- Methylated circulated tumor DNA (meth-ctDNA) has potential as a universal biomarker
- Patients with pathological lymph node involvement could not be identified using a multiplex DNA methylation analysis of tumor-specific and organ-specific markers
- Preoperative multiplex methylation analysis of tumor specific and organ specific markers in colon cancer holds prognostic value
- Preoperative positive meth-ctDNA could represent a biomarker for high-risk patients in the risk classification of colon cancer. Randomized trials investigating the efficacy of NAC in colon cancer patients with preoperative positive meth-ctDNA are suggested.

## Acknowledgements

The authors would like to thank the study participants and the staff at Vejle Hospital who collected samples for this study. The authors are thankful for the funding received from The Moltum Foundation, The Grønbeck-Olsen Foundation and The Højmosegård Foundation. We would like to acknowledge the support from OPEN, Open Patient data Explorative Network, Odense University Hospital, Region of Southern Denmark. Study data were managed using REDCap (electronic data capture tool) hosted by OPEN. We express gratitude to Karin Larsen for linguistic editing of the manuscript.

## Ethics approval

The study was conducted in accordance with the Declaration of Helsinki. The study was approved by the Regional Committee on Health Research Ethics for Southern Denmark (S-20190181).

## Informed consent

Written and orally informed consent to translational research according to the Helsinki II Declaration was obtained from all patients.

## Data Availability Statement

The datasets analyzed during the current study are available from the corresponding author upon reasonable request.

## Funding

The study received funding from The Moltum Foundation, The Grønbeck-Olsen Foundation and The Højmosegård Foundation. The study was financially supported by the Department of Oncology, Vejle Hospital, University Hospital of Southern Denmark. No financial support was given to any of the authors.

## Disclosure statement

The authors declare no competing interests.

## Author contributions

Conzeptualization, LR, LHJ, RFA, TFH; Methodology, RFA, MMJ; Validation, RFA; Data curation, LR, JL, SR, TFH, LHJ; Formal analysis, ST, LR; Investigation, LR; Funding acquisition, LR; Supervision, LHJ, RFA, TFH; Writing – original draft: LR; Writing – review and editing: LR, LHJ, RFA, TFH, ST, MMJ, JL, SR. All authors read and approved the final manuscript.

## Notes

### Competing Interest Statement

The authors have declared no competing interest.

### Funding Statement

The study received funding from The Moltum Foundation, The Groenbeck-Olsen Foundation and The Hoejmosegaard Foundation. The study was financially supported by the Department of Oncology, Vejle Hospital, University Hospital of Southern Denmark.

### Author Declarations

The Regional Committee on Health Research Ethics for Southern Denmark gave ethical approval for this work

