## Supplemental Figure 1 for "Development and clinical application of a methylated ctDNA assay in the preoperative risk classification of resectable colon cancer"

**Supplemental Figure 1** Positive and negative plasma samples in the multiplex analysis.

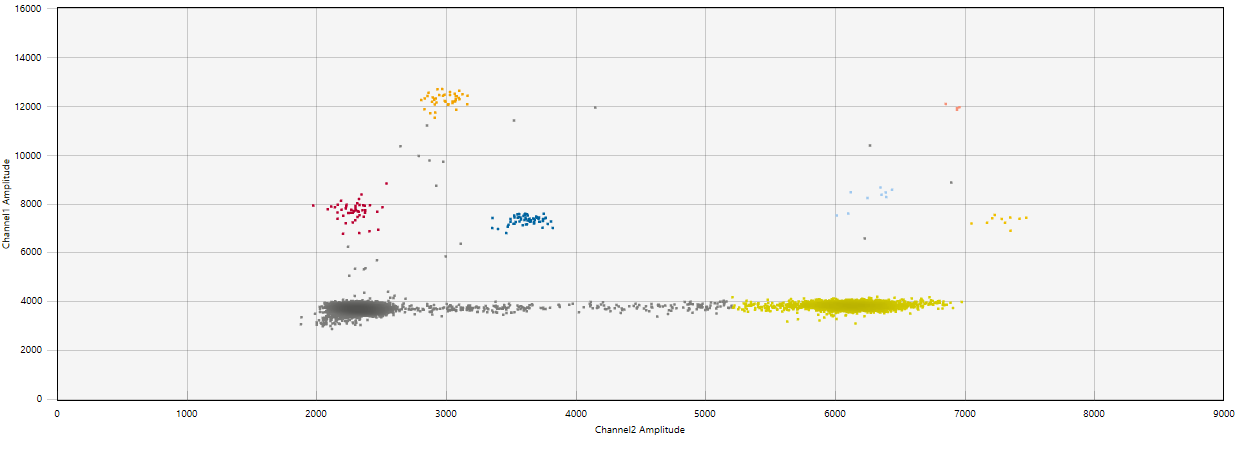

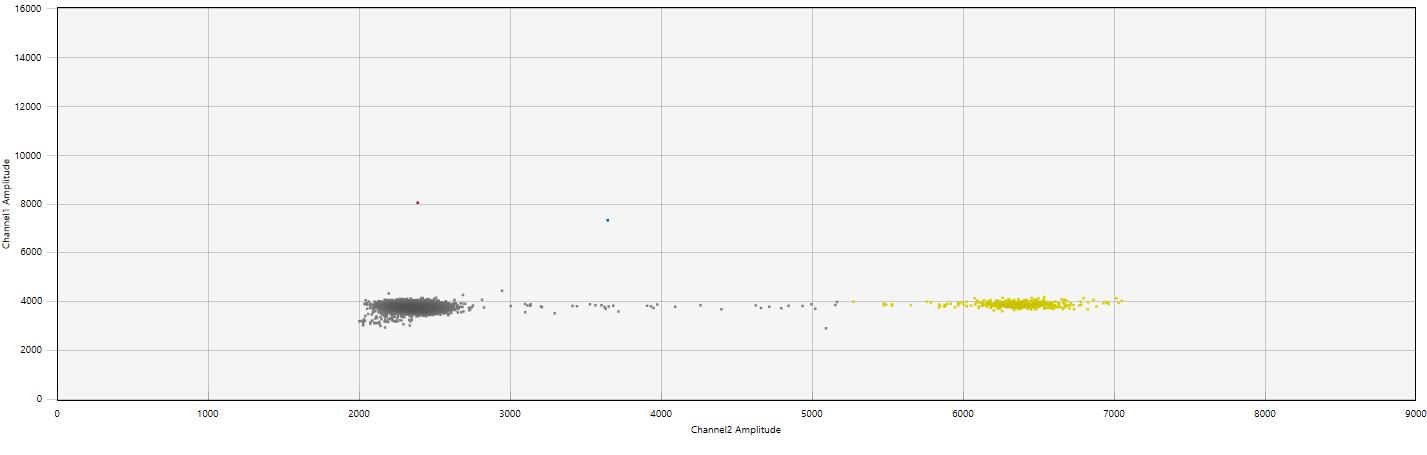

 Red, NPY; Yellow, KANK1 as; Blue, GAL3ST3; Light yellow, ALB
