## Supplemental Figure 2 for "Development and clinical application of a methylated ctDNA assay in the preoperative risk classification of resectable colon cancer"

**Supplemental Figure 2** (a-d) Percentage of the three meth-ctDNA markers measured by singleplex and multiplex analysis in colon and rectal tumor tissue; (e) Percentage of the three meth-ctDNA markers measured by multiplex analysis in colon and rectal normal tissue

**2a** KANK1 (KN Motif And Ankyrin Repeat Domains 1) measured by singleplex

KANK1

% methylated

|  | KANK1 single | KANK1 multi |
| --- | --- | --- |
| Colon T1 | 28.951 | 26.2838 |
| Colon T2 | 24.6419 | 25.6836 |
| Colon T3 | 22.898 | 21.8807 |
| Colon T4 | 3.38961 | 2.41803 |
| Colon T5 | 31.0791 | 29.5652 |
| Colon T6 | 28.5903 | 29.6854 |
| Colon T7 | 20.479 | 23.0337 |
| Colon T8 | 5.10417 | 5.89041 |
| Colon T9 | 39.9371 | 38.3117 |
| Colon T10 | 23.5974 | 20.9967 |
| Rectum T1 | 20.5528 | 15.7325 |
| Rectum T2 | 18.32 | 20.5263 |
| Rectum T3 | 7.72973 | 6.94611 |
| Rectum T4 | 15.9643 | 14.0511 |
| Rectum T5 | 14.4526 | 13.9163 |
| Rectum T6 | 7.95122 | 8.35897 |
| Rectum T7 | 23.5135 | 26.6443 |
| Rectum T8 | 27.1287 | 24.1697 |
| Rectum T9 | 25.1781 | 30.1546 |
| Rectum T10 | 17.451 | 15.6954 |

**2b** GAL3ST3 (Galactose-3-O-Sulfotransferase 3) measured by singleplex

GAL3ST3

% methylated

|  | GAL3ST3 single | GAL3ST3 multi |
| --- | --- | --- |
| Colon T1 | 55.4307 | 54.3919 |
| Colon T2 | 35.3933 | 34.5845 |
| Colon T3 | 27.881 | 26.4014 |
| Colon T4 | 2.44872 | 2.41803 |
| Colon T5 | 33 | 27.8261 |
| Colon T6 | 42.9919 | 42.9549 |
| Colon T7 | 26.9942 | 30.618 |
| Colon T8 | 7.16312 | 6.64384 |
| Colon T9 | 47.7966 | 44.4805 |
| Colon T10 | 24.7651 | 25.3821 |
| Rectum T1 | 23.1746 | 24.1401 |
| Rectum T2 | 35.7143 | 38.4211 |
| Rectum T3 | 24.9708 | 28.024 |
| Rectum T4 | 19.8932 | 19.8905 |
| Rectum T5 | 17.6978 | 19.8479 |
| Rectum T6 | 12.25 | 11.9487 |
| Rectum T7 | 30.687 | 33.8255 |
| Rectum T8 | 33.0693 | 28.2288 |
| Rectum T9 | 38.5 | 41.2371 |
| Rectum T10 | 17.6389 | 17.0861 |

**2c** NPY (Neuropeptide Y) measured by singleplex

NPY

% methylated

|  | NPY single | NPY multi |
| --- | --- | --- |
| Colon T1 | 52.5362 | 53.3784 |
| Colon T2 | 16.3764 | 17.5871 |
| Colon T3 | 18.2482 | 20.9765 |
| Colon T4 | 1.85185 | 3.45492 |
| Colon T5 | 20.1439 | 17.5362 |
| Colon T6 | 25.1682 | 25.0342 |
| Colon T7 | 19.0751 | 19.2697 |
| Colon T8 | 6.01351 | 5.23973 |
| Colon T9 | 35.5769 | 43.1818 |
| Colon T10 | 47.619 | 49.5017 |
| Rectum T1 | 10.4188 | 13.758 |
| Rectum T2 | 32.6695 | 37.807 |
| Rectum T3 | 21.1364 | 22.0359 |
| Rectum T4 | 18.1618 | 15.292 |
| Rectum T5 | 21.1896 | 28.5932 |
| Rectum T6 | 6.31841 | 7.4359 |
| Rectum T7 | 25.9627 | 27.651 |
| Rectum T8 | 17.264 | 16.3284 |
| Rectum T9 | 44.2584 | 52.5773 |
| Rectum T10 | 18.6111 | 15.2318 |

**2d** All three meth-ctDNAs measured by multiplex in colon and rectal tumor tissue

% methylated

|  | KANK1 multi | GAL3ST3 multi | NPY multi |
| --- | --- | --- | --- |
| Colon T1 | 26.2838 | 54.3919 | 53.3784 |
| Colon T2 | 25.6836 | 34.5845 | 17.5871 |
| Colon T3 | 21.8807 | 26.4014 | 20.9765 |
| Colon T4 | 2.41803 | 2.41803 | 3.45492 |
| Colon T5 | 29.5652 | 27.8261 | 17.5362 |
| Colon T6 | 29.6854 | 42.9549 | 25.0342 |
| Colon T7 | 23.0337 | 30.618 | 19.2697 |
| Colon T8 | 5.89041 | 6.64384 | 5.23973 |
| Colon T9 | 38.3117 | 44.4805 | 43.1818 |
| Colon T10 | 20.9967 | 25.3821 | 49.5017 |
| Rectum T1 | 15.7325 | 24.1401 | 13.758 |
| Rectum T2 | 20.5263 | 38.4211 | 37.807 |
| Rectum T3 | 6.94611 | 28.024 | 22.0359 |
| Rectum T4 | 14.0511 | 19.8905 | 15.292 |
| Rectum T5 | 13.9163 | 19.8479 | 28.5932 |
| Rectum T6 | 8.35897 | 11.9487 | 7.4359 |
| Rectum T7 | 26.6443 | 33.8255 | 27.651 |
| Rectum T8 | 24.1697 | 28.2288 | 16.3284 |
| Rectum T9 | 30.1546 | 41.2371 | 52.5773 |
| Rectum T10 | 15.6954 | 17.0861 | 15.2318 |

**2e** All three meth-ctDNAs measured by multiplex in colon and rectal normal tissue

% methylated

|  | KANK1 multi | GAL3ST3 multi | NPY multi |
| --- | --- | --- | --- |
| Colon N1 | 14.7083 | 6.10417 | 1.38333 |
| Colon N2 | 19.0529 | 15.7489 | 3.89868 |
| Colon N3 | 8.77612 | 0.855224 | 0.191045 |
| Colon N4 | 17.0229 | 2.0687 | 1.35496 |
| Colon N5 | 19.1827 | 11.0577 | 2.40385 |
| Colon N6 | 20.9482 | 6.90187 | 1.02536 |
| Colon N7 | 10.5308 | 3.47407 | 0.554885 |
| Colon N8 | 24.1935 | 4.1129 | 1.12298 |
| Colon N9 | 16.0377 | 2.55765 | 0.285115 |
| Colon N10 | 17.5547 | 4.83577 | 0.600365 |
| Rectum N1 | 22.2549 | 11.6667 | 2.44118 |
| Rectum N2 | 23.0556 | 11.8056 | 3.40972 |
| Rectum N3 | 23.2344 | 15.0461 | 1.53531 |
| Rectum N4 | 20.5263 | 7.69737 | 1.52632 |
| Rectum N5 | 25.4867 | 12.6549 | 0.861062 |
| Rectum N6 | 22.8099 | 24.2975 | 3.47107 |
| Rectum N7 | 22.4691 | 4.0679 | 0.802469 |
| Rectum N8 | 21.8617 | 13.1915 | 1.05851 |
| Rectum N9 | 23.3333 | 12.5786 | 1.16352 |
| Rectum N10 | 21.2963 | 18.3128 | 2.34568 |
