## Supplemental Figure 3 for "Development and clinical application of a methylated ctDNA assay in the preoperative risk classification of resectable colon cancer"

Excluded, N=57

- Neoadjuvant chemotherapy, N=9
- Blood sample drawn post-surgery, N=25
- Other malignant disease within 3 years before surgery, N=20
- Synchronous metastatic disease, N=7

Randomization

Mkldæfshnfjksdn fsdkæ

mkfldsnklsdnfkls

Allocated to validation cohort

N=101

Allocated to discovery cohort

N=102

Inclusion

N=203

Assessment for enrollment

N=260

**Flow chart**
