## Supplemental Figure 4 for "Development and clinical application of a methylated ctDNA assay in the preoperative risk classification of resectable colon cancer"

**Supplemental Figure 4** Receiver operating characteristic (ROC) curves of the ability of preoperative meth-ctDNA to predict postoperative lymph node metastases (pN+) in the discovery cohort (a-f) and the validation cohort (g-l**)**

**Discovery cohort**

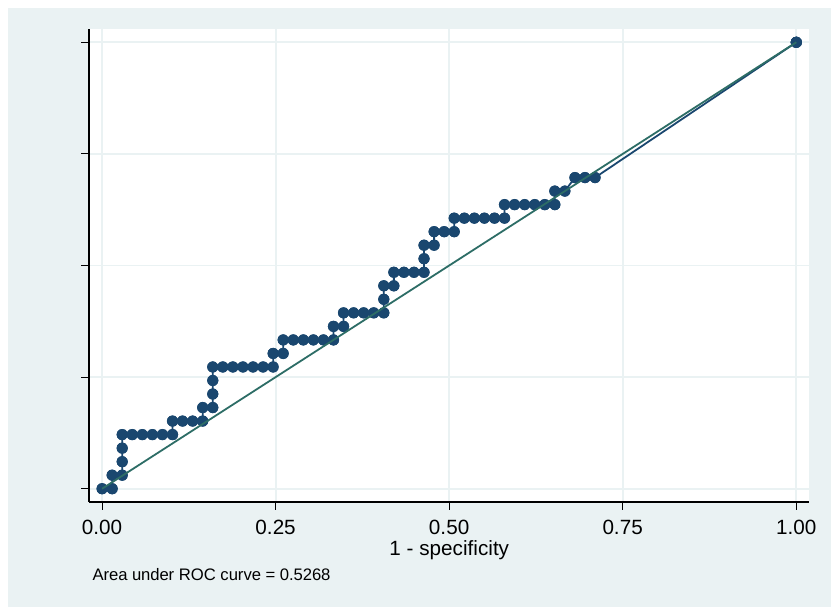

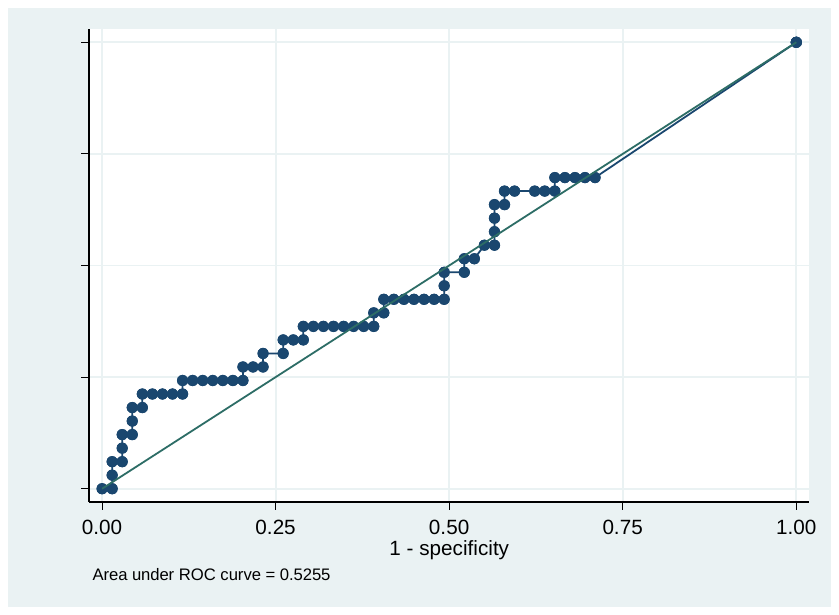

4a Mean percentage meth-ctDNA predicting pN+, AUC 53% 4b Mean copies meth-ctDNA predicting pN+, AUC 53%

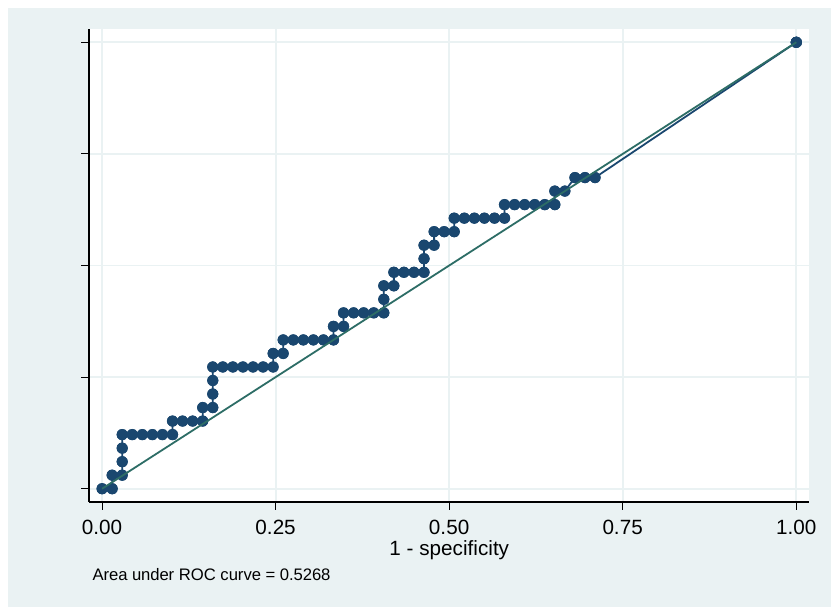

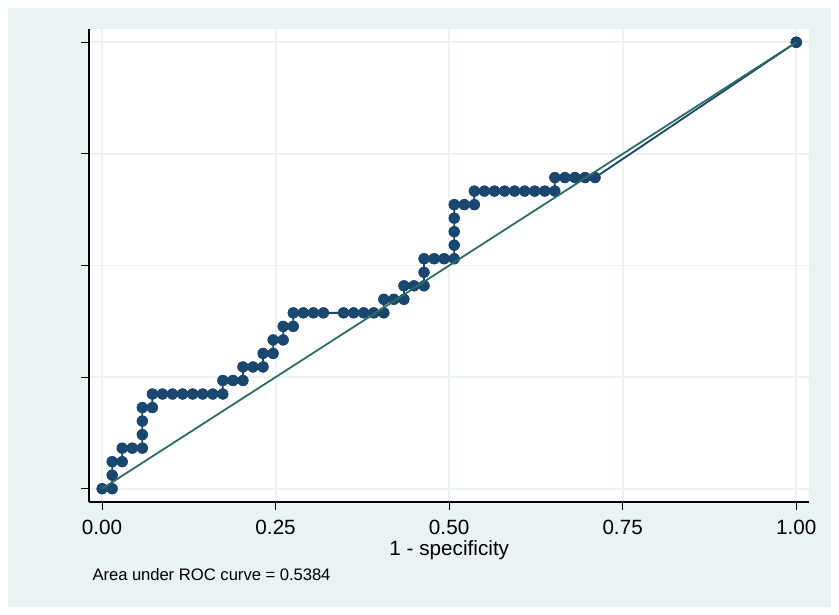

4c Sum percentage meth-ctDNA predicting pN+, AUC 53%	4d Sum copies meth-ctDNA predicting pN+, AUC 54%

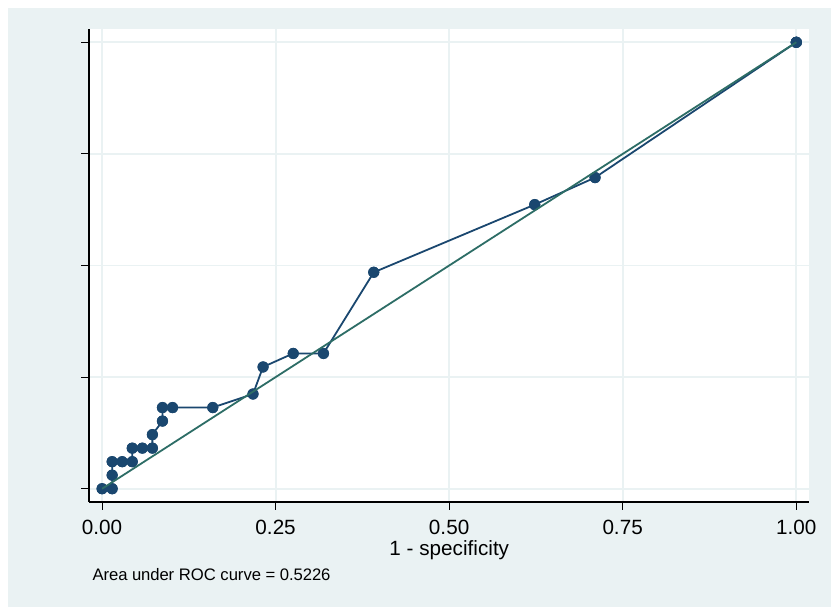

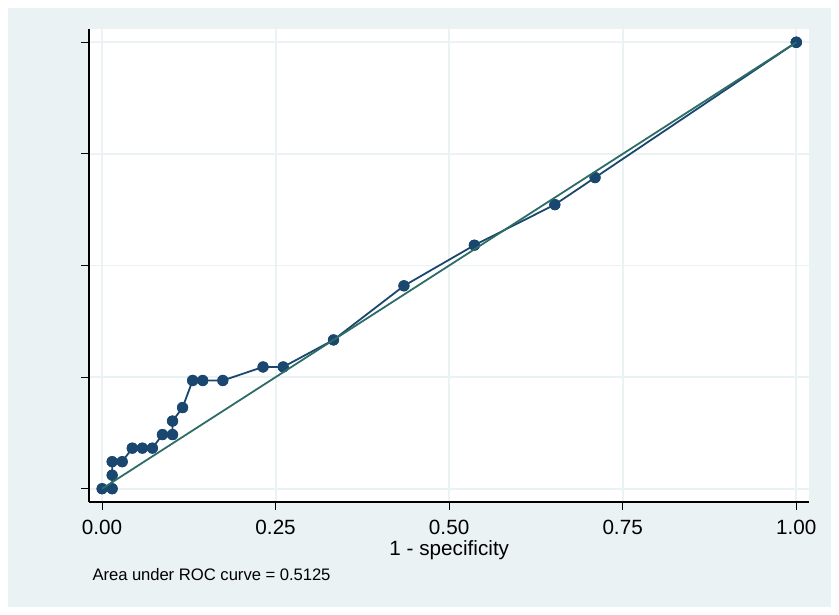

4e Max percentage meth-ctDNA predicting pN+, AUC 53%	4f Max copies meth-ctDNA predicting pN+, AUC 51%

**Validation cohort**

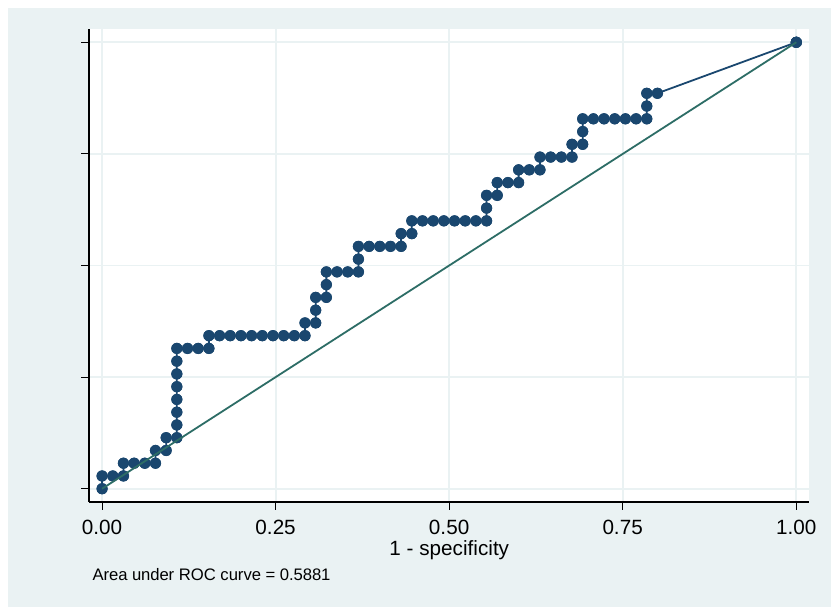

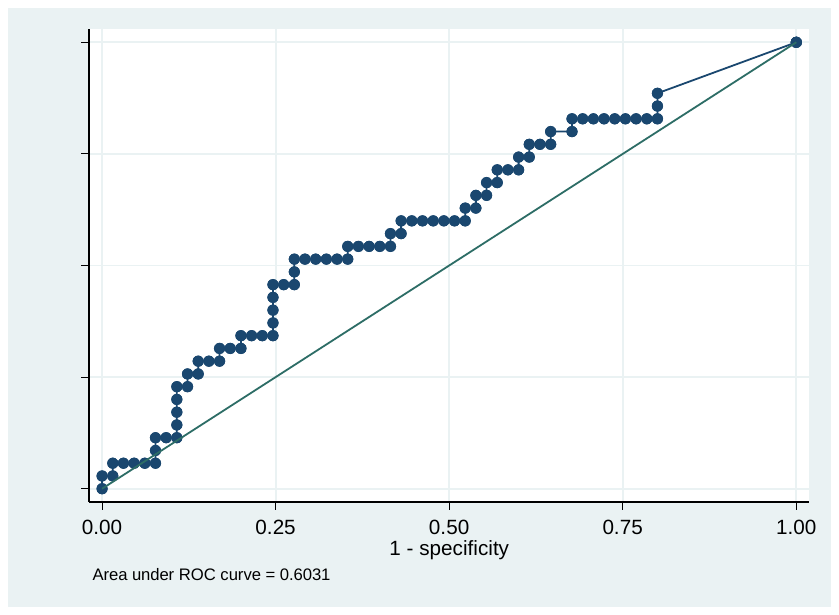

4g Mean percentage meth-ctDNA predicting pN+, AUC 59% 4h Mean copies meth-ctDNA predicting pN+, AUC 60%

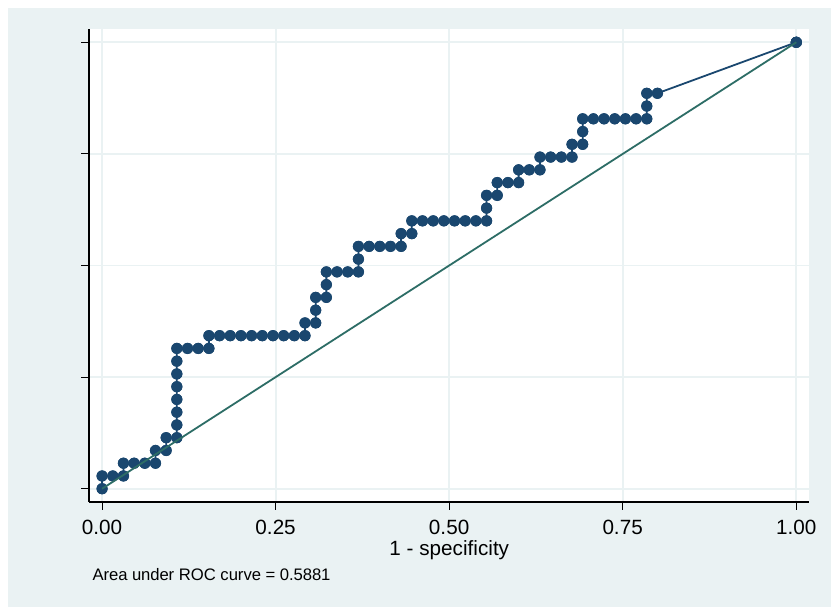

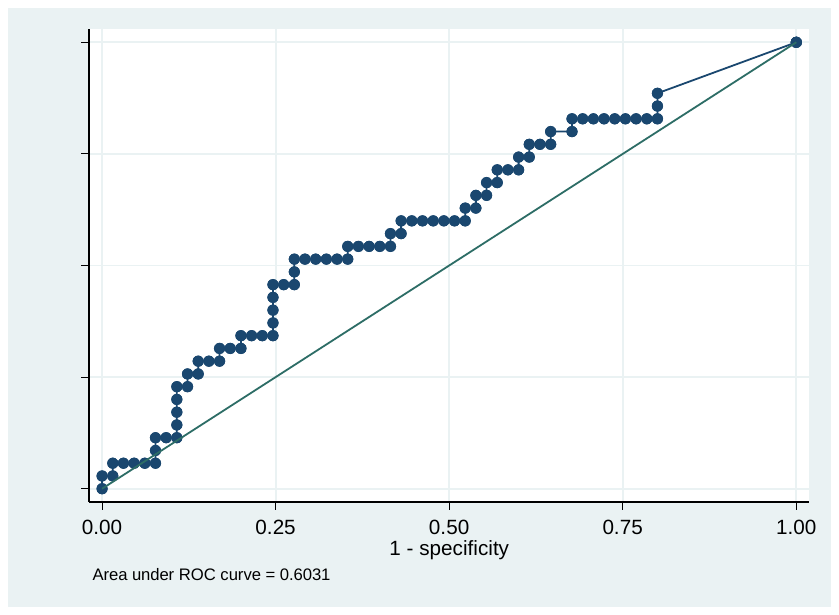

4i Sum percentage meth-ctDNA predicting pN+, AUC 59%	4j Sum copies meth-ctDNA predicting pN+, AUC 60%

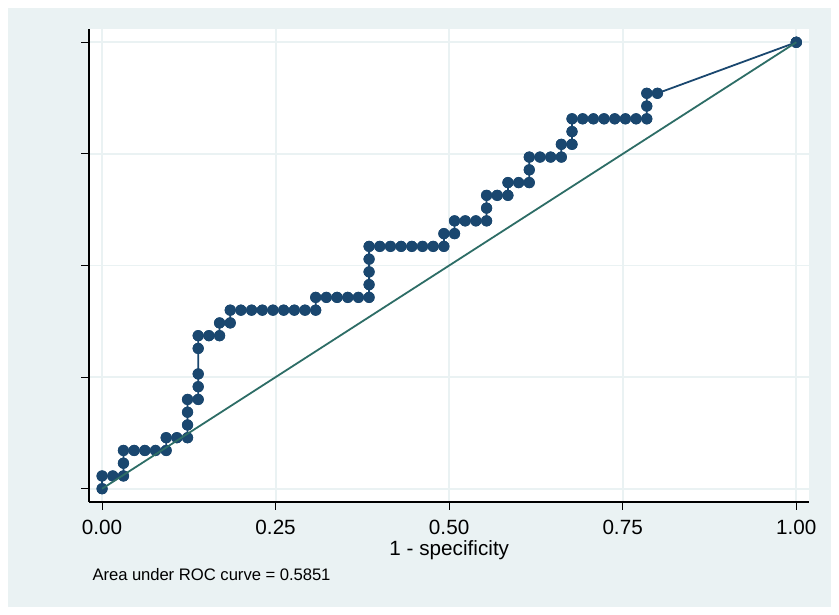

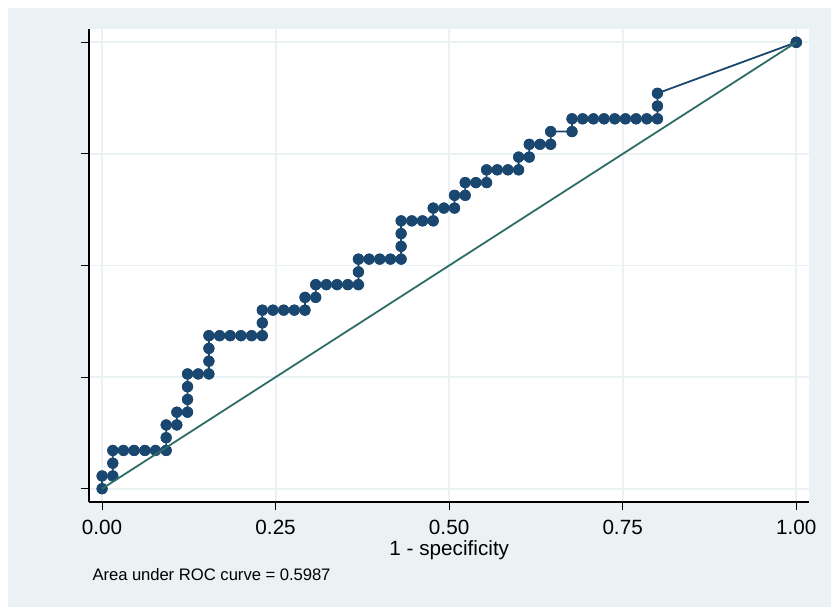

4k Max percentage meth-ctDNA predicting pN+, AUC 58%	4l Max copies meth-ctDNA predicting pN+, AUC 59%
