## Supplemental Table 2 for "Development and clinical application of a methylated ctDNA assay in the preoperative risk classification of resectable colon cancer"

**Supplemental Table 2** Colon multiplex assay describing sample and mastermix combinations (a+b) and details on primers and probes (c)

**2a**

| Supermix for probes 2x | 24 µl | |
| --- | --- | --- |
| Colon multiplex mix 5x | 10 µl | |
| H2O | 2 µl | |
| Sample | 12 µl | |
| Total | 48 µl | |
| **2b** | |  |
| **5x colon multiplex mix** | |  |
| NPY FAM 40x assay | | 1.50 |
| KANK1 as FAM 40x assay | | 1.50 |
| GAL3ST3 FAM 40x assay | | 1.00 |
| GAL3ST3 HEX 40x assay | | 0.42 |
| ALB 10x assay | | 5.00 |
| H2O | | 0.58 |
| Total | | 10.00 |

NPY, Neuropeptide Y; KANK1, KN Motif And Ankyrin Repeat Domains 1; GAL3ST3, Galactose-3-O-Sulfotransferase 3; ALB, albumin; as, antisense

| **2c**  Assay | Name | | Sequence | | Manufacturer |
| --- | --- | --- | --- | --- | --- |
| NPY [12] | NPY F | | CGCGGCGAGGAAGTTTTATA | | LGC Biosearch |
|  | NPY R | | ATACTATCGAACGAACGTCTCCG | | LGC Biosearch |
|  | NPY Probe | | FAM-CGCGATTCGTTTTTTGTA-IOWA | | IDT |
| KANK1 as | KANK1 as F | | AATTCGGTTAGCGGTGGTC | | Biomers |
|  | KANK1 as R | | GAAACAACGCGAACTAACGAAA | | Biomers |
|  | KANK1 as Probe | | FAM –ATTTATTACGTGGCGTTCGTCGCG-BHQ1 | | Biomers |
| GAL3ST3 | GAL3ST3 F | | TTTGCGCGTCGAGTGTC | | Biomers |
|  | GAL3ST3 R | | AAACGAACCTAACGAACGCT | | Biomers |
|  | GAL3ST3 Probe | | FAM-AGCGTTTGGATGGGATTAGGTGGT-IOWA | | IDT |
| GAL3ST3 | GAL3ST3 F | | TTTGCGCGTCGAGTGTC | | Biomers |
|  | GAL3ST3 R | | AAACGAACCTAACGAACGCT | | Biomers |
|  | GAL3ST3 Probe | | HEX-AGCGTTTGGATGGGATTAGGTGGT-IOWA | | IDT |
| ALB [12] | ALB F | | GGGATGGAAAGAATTTTATGTT | | LGC Biosearch |
|  | ALB R | | AAACAAACTAACCCCAAATTCT | | LGC Biosearch |
|  | ALB Probe | | VIC-AGGGTTTTTATAATTTA-MGBNFQ | | Thermo Fisher |

NPY, Neuropeptide Y; KANK1, KN Motif And Ankyrin Repeat Domains 1; GAL3ST3, Galactose-3-O-Sulfotransferase 3; ALB, albumin; as, antisense; F, forward; R, reverse
